# The Feasibility of Studying Metabolites in PICU Multi-Organ Dysfunction Syndrome Patients Over an 8-day Course Using An Untargeted Approach

**DOI:** 10.1101/2020.12.04.20244053

**Authors:** Mara L. Leimanis-Laurens, Danny Gil, Andrew Kampfshulte, Claire Krohn, Elizabeth Prentice, Dominic Sanfilippo, Jeremy W. Prokop, Todd Lydic, Surender Rajasekaran

## Abstract

Metabolites are generated from critical biological functions and metabolism. This pediatric study reviewed plasma metabolites in patients suffering from multi-organ dysfunction syndrome (MODS) in the pediatric intensive care unit (PICU) using an untargeted metabolomics approach. Patients meeting criteria for MODS were screened for eligibility and consented (n=24), and blood samples were collected at baseline, 72 hours, and 8 days; control patients (n=4), were presenting for routine sedation in an outpatient setting. A sub-set of MODS patients (n=8) required additional support with veno-atrial extracorporeal membrane oxygenation (VA-ECMO) therapy. Metabolites from thawed blood plasma were determined from ion pairing reversed-phase LC-MS analysis. Chromatographic peak alignment, identification, relative quantitation, statistical and bioinformatics evaluation were performed using MAVEN and MetaboAnalyst 4.0. Metabolite analysis revealed 115 peaks per sample. From the PLS-DA with VIP scores above ≥2.0, 7 dynamic metabolites emerged over the 3 time points: tauro-chenodeoxycholic acid (TCDCA), hexose, *p*-hydroxybenzoate, hydroxyphenylacetic acid (HPLA), 2_3-dihydroxybenzoic acid, 2-keto-isovalerate, and deoxyribose phosphate. After Bonferonni adjustment for repeated measures hexose and *p*-hydroxybenzoate were significant at one time point, or more. Kendall’s tau-b test was used for internal validation of creatinine. Metabolites may be benign or significant in describing a patient’s pathophysiology and require operator interpretation.

## 1. Introduction

The ability to identify, quantify, and analyze the metabolic profile of a pediatric patient, allows us to investigate the interaction between both physiologic and pathologic states. Metabolites are under the control of environmental pressures, such as nutrition [1], viral infections (such as Covid-19 [2], Ebola [3]), gut bacterial composition and cancer [4], medications [5,6], and a patient’s own pre-existing genetic make-up [7]. Metabolites, being low molecular weight molecules and/or products of metabolic pathways, are growing in appeal medically over the last decade for their potential in disease characterization, drug discovery and precision medicine [8,9].

We have previously described the current cohort of patients for patient whole blood transcriptomics [10,11], and plasma lipidome [12]. This has revealed a complex biology in a heterogenous patient population with a non-uniform patient response to treatments over an 8-day course (stabilization and recovery phases) of illness during a PICU admission. Complimentary to these previously reported analytic modalities from whole blood [10-12], the aim of this current report was two-fold: 1) to characterize total blood plasma metabolites (polar, charged) using an untargeted approach, 2) to determine change in metabolites over an 8-day PICU course. There is a gap in our understanding of the complex interaction between pediatric critical illness, specifically multi-organ dysfunction syndrome (MODS) [13] (affecting twenty percent of PICU admissions [14], resulting in ten times the mortality rate [15]), and their respective blood metabolites.

## 2. Materials and Methods

### 2.1 Study Population, Site and Sample Collection

After IRB approval, a short-term longitudinal design was adopted at Helen DeVos Children’s Hospital (2016-062-SH/HDVCH). Samples were collected under the protocol and study design [10-12] in a quaternary-care, urban, pediatric hospital in Western, Michigan. In brief, patients who were identified as having MODS were enrolled, 24 in total, with an additional 4 sedation-control patients. These 24 patients were then further classified as needing veno-arterial extracorporeal membrane oxygenation (VA-ECMO) as a therapeutic modality (n=8) according to Extracorporeal Life Support Organization (ELSO) criteria [16]. Blood samples from the patients were obtained and placed into EDTA-filled tubes, plasma was processed and stored at −80 °C for later use. All samples had undergone one freeze-thaw before processing and analysis.

### 2.2 Metabolite Extraction and Liquid Chromatography-Mass Spectrometry (LC-MS)

Plasma samples (∼50 microliters) were subjected to biphasic extraction using chloroform/methanol/water as described previously [17] to remove nonpolar matrix interferences and recover polar metabolites in the aqueous extraction phase. Stable isotope labeled (D^4^)-succinate was added to plasma during extraction for use in estimation of metabolite recovery and for relative quantitation across experimental groups. Samples were filtered through 0.2 micron syringe filters (Fisher Scientific) and reconstituted in 100 microliters of 50 % methanol for use in ion pairing reversed-phase LC-MS analysis.

Targeted polar metabolite identification utilized a Thermo Scientific model TSQ Vantage triple quadrupole mass spectrometer operating in negative ion mode. The mass spectrometer was coupled to a Shimadzu Prominence HPLC with thermostated column oven and autosampler. Ten microliter sample injections were subjected to gradient elution with (A) 10 mM tributylamine and 15 mM acetic acid (pH 4.95), and (B) Methanol according to B. Luo, et al. [18], with separation of metabolites achieved on a Phenomenex Synergi Hydro-RP C18 column (2.0 mm x 150 mm, 3 micron particles, 80 Angstrom pore size). The column was protected by a Phenomenex guard cartridge of identical chemistry. Metabolites were identified and quantitated by selected ion monitoring. Detection parameters for each precursor/product ion pair of interest have been optimized based using commercially available standards.

### 2.3 Data analysis

LC-MS data analysis of chromatographic peak alignment, compound identification, relative quantitation, and statistical evaluation across experimental groups will be performed using MAVEN software [19]. Only relative quantitation of analytes against a selected internal standard was performed for comparison of values across experimental treatment groups. “Absolute” quantitation was not carried out in these experiments. Metabolites as listed in Supplemental Table 1, were initially categorized as anabolic vs. catabolic and endogenous vs. exogenous according to Human Metabolite Data Base (HMDB) (https://hmdb.ca/metabolites/) (Supplemental Figure 1).

Metabolic profiles for sedation-controls were compared to MODS or ECMO patients, quantified as percent of total. Metabolites with >30% of cases with zero values were excluded from further analysis; consequently 66 metabolites were analyzed over the three time points (Figure 1). Using MetaboAnalyst 4.0 [20], data were normalized using pareto scaling (mean-centered and divided by the square root of standard deviation of each variable), and subjected to a multi-variate partial least squares-discriminant analysis (PLS-DA) analysis [21], using Q2 values for cross-validation [22]. No data points were excluded. To assess the significance of class discrimination, a permutation test was performed and PLS-DA model built [between the data (X) and the permuted class labels (Y)] using the optimal number of components as determined by cross validation for the model based on the original class assignment [23]. This guided analysis allowed for the display of each specific group assignment. Variable Importance in Projection (VIP) is a weighted sum of squares of the PLS loadings considering the amount of explained Y-variation in each dimension.

**Figure 1:**
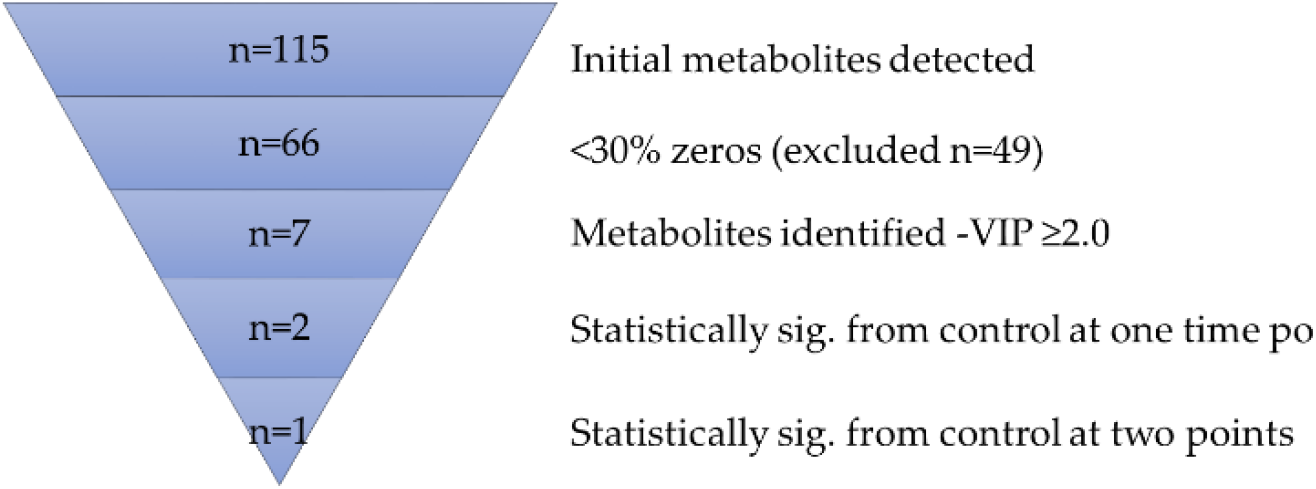
Overview-untargeted analysis

Univariate analysis was performed using MedCalc (MedCalc Software Ltd, Ostend, Belgium) for candidate metabolites as determined by variance of importance (VIP) scores >2.0 from seven metabolites, using independent T-tests (equal variances), and Welch-test (unequal variances). A Bonferroni correction of P-value (<0.008) was used to identify statistically significant associations with metabolites (to control for Type I errors), which was calculated by dividing the significance threshold of 0.05 by the number of repeated measures; in this case MODS and ECMO compared to sedation-controls (at baseline, time 72hrs and 8 days). Box and whisker plots were generated for the two remaining metabolites of interest, which included the median, the interquartile range (box), the outer range (whiskers) to pictorially summarize the central tendency, dispersion, skewness, and extremes of the dataset [24].

## 3. Results

### 3.1 Metabolite Ontology and Origin

All 115 metabolites are listed in Supplemental Table 1 according to compound identification from the metabolic mass to charge ratio and retention time. From here we looked at the ontology of the metabolites in order to characterize according to any known function and origin. We found that the majority of metabolites detected were endogenous in nature (76%), and associated with catabolic (38%), anabolic (35%), both catabolic and anabolic metabolism (14%), or unspecified mechanisms of action (13%) (Supplement Figure 1 A & B), according to HMDB (https://hmdb.ca/metabolites/).

### 3.2 Bioinformatic Analysis

Analytical flow chart is presented in Figure 1. Percent total of metabolites and change over time were visualized using supervised analysis PLS-DA, which revealed clustering of ECMO patients within the MODS patients over the three time points, as compared to the sedation-control group (Figure 2, A, B, C). These groupings were also visualized by heatmap analysis (Supplemental Figure 2, A, B, C), supporting the initial finding of the PLS-DA, whereby the sedation-control patients were found to cluster amongst themselves.

**Figure 2:**
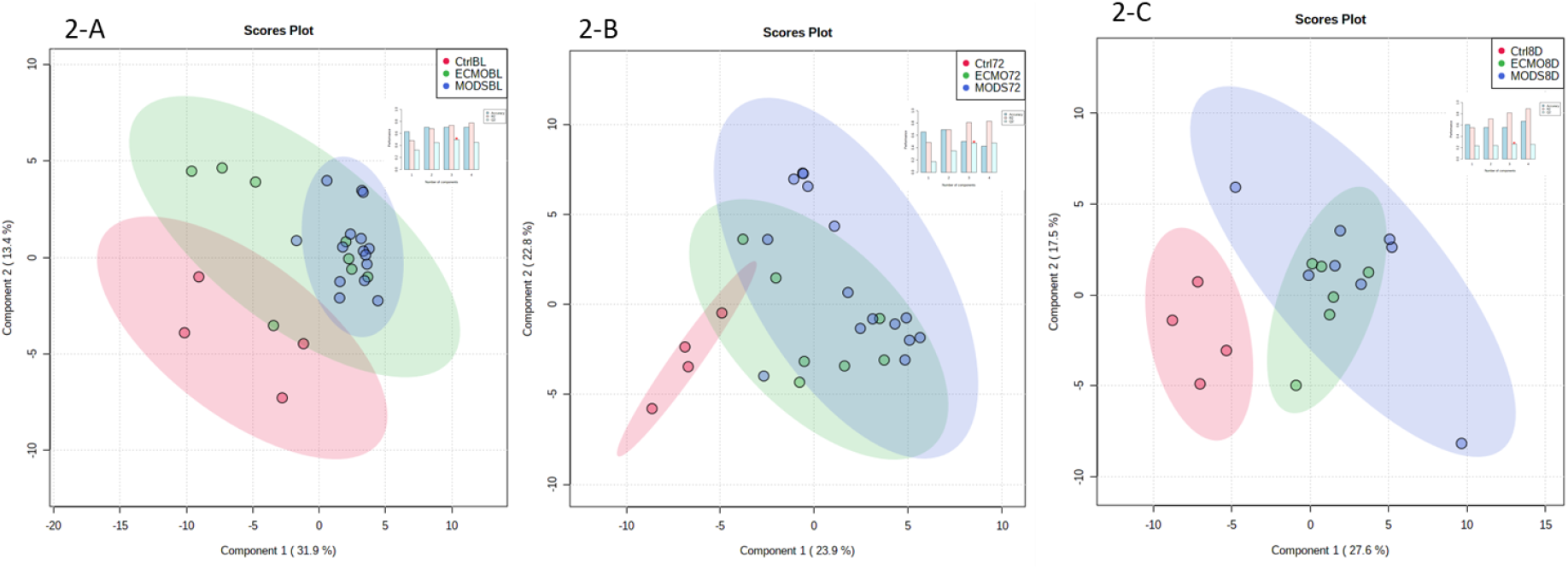
PLS-DA plot of all patient samples at baseline (A), 72 hours (B), and 8 days (C) Insert: PLS-DA classification using different number of components. The red star indicates the best classier.

The important features as identifies by the PLS-DA at baseline (Figure 3A), at 72 hours (Figure 3B) and at 8 days (Figure 3C), reveal seven metabolites with relative concentrations of the corresponding metabolite in each group under study. In total seven metabolites of interest emerged over the 3 time points: tauro-chenodeoxycholic acid (TCDCA)-*a conjugated bile acid*, hexose-monosaccharide-*simple sugar, p*-hydroxybenzoate-*biocide-antimicrobial agent/tyrosine, tryptophan, phenylalanine metabolite*, hydroxyphenylacetic acid (HPLA)-*metabolite of phenylalanine*, 2_3- dihydroxybenzoic acid-*drug metabolite*, 2-keto-isovalerate-*cellular intermediate for the synthesis of branched-chain amino acids*, deoxyribose phosphate-*a pentose phosphate*.

**Figure 3:**
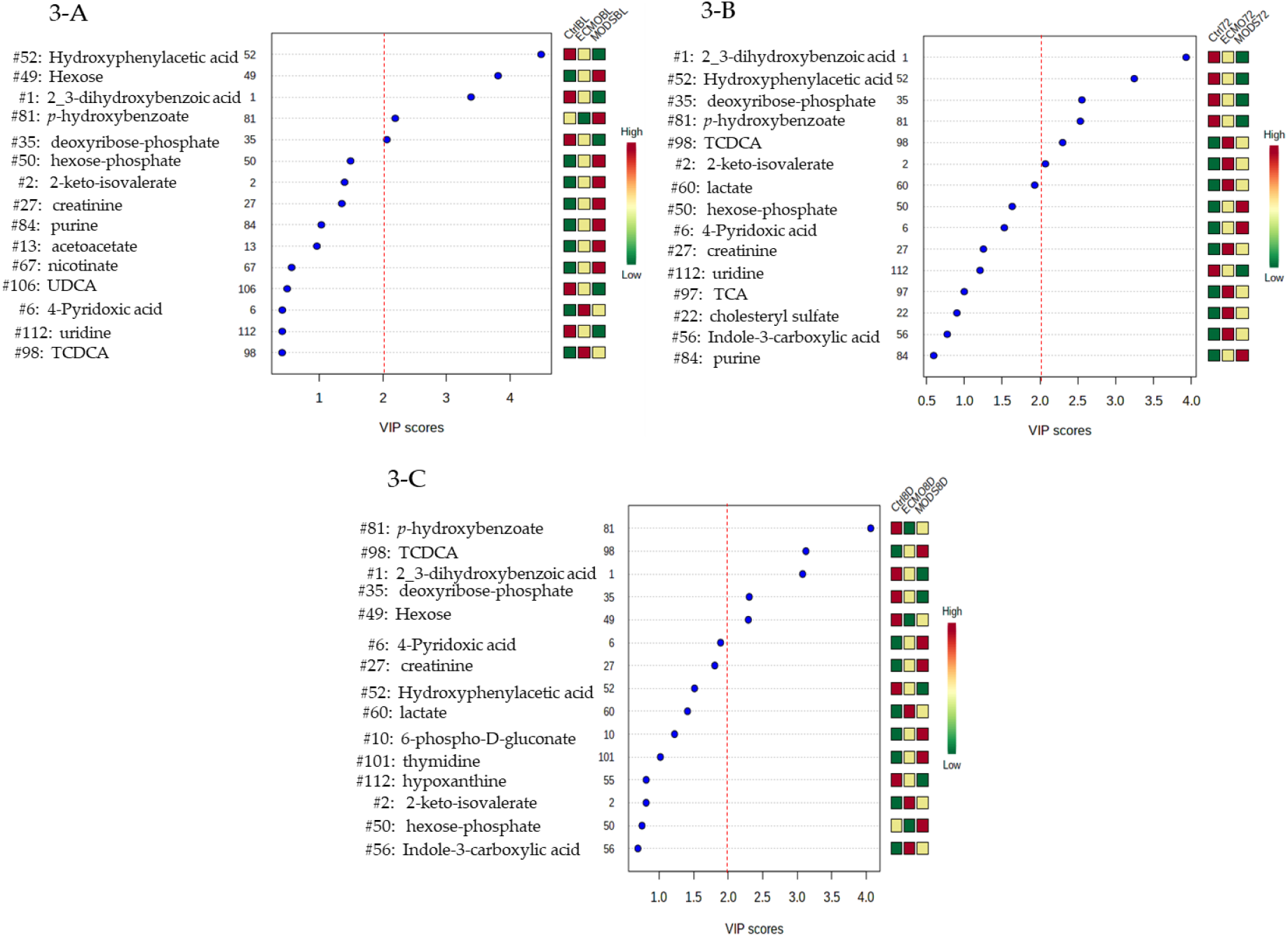
Important features identified by PLS-DA at baseline (A), 72 hours (B), and 8 days (C). The colored boxes on the right indicate the relative concentrations of the corresponding metabolite in each group under study. TCDCA: tauro-chenodeoxycholic acid.

Starting with HPLA (with a VIP score ≥1.5 at 8 days) over the 3-time points, sedation-control values are consistently highest, with ECMO patients demonstrating an intermediate profile and MODS patients with the lowest values according to the relative concentrations. 2_3- dihydroxybenzoic acid and deoxyribose-phosphate share similar relative concentration profiles to HPLA with the values highest for sedation-control patients compared to MODS with the lowest relative concentrations.

In the exact opposite profile is hexose, which reveals the highest relative concentrations values in MODS patients as compared to sedation-controls, again with ECMO sharing intermediate profiles at baseline. By day 8 however, this has changed completely, whereby sedation-controls have the highest relative concentrations, and ECMO the lowest, with MODS demonstrating intermediate profiles. We may extrapolate from this that the patients with critical illness demonstrate some fluctuations in hexose over time, and that the detection of this blood plasma metabolite is a dynamic process. Patients were neither hyper- nor hypo-glycemic according to their clinical glucose levels (also a 6-carbon sugar-*data not shown*), and this is closely monitored at the PICU bedside, given blood glucose levels have been previously demonstrated to adversely affect patient outcomes, especially in the case of hyperglycemia [25].

Remaining metabolites of interest include *p*-hydroxybenzoate with exception of the 72 hours’ time point, reveal that ECMO patients have lower relative concentrations than MODS patients, and TCDCA, which is consistently lower in sedation-control patients, as compared to both MODS and ECMO. TCDCA is a maker of liver injury, which is still elevated after 8 days for MODS and ECMO patients. It is believed that shock liver common in this patient population usually subsides after a few days, from our results we may speculate that this metabolite is still present at the 8^th^ day post-study enrollment. Lastly, keto-isovalerate like hexose and TCDCA has higher relative concentrations for both MODS and ECMO patients compared to sedation-controls at both baseline and 72 hours (at 8-days similar patterning, however VIP score <1.0). Additional metabolites of note include lactate (VIP score ≥1.5) which would be expected in this group of patients [24], which is highest in ECMO patients, second in MODS and lowest in the sedation-controls at the 72-hour time point.

### 3.3 Repeated Measures Over 3 Time Points

Furthermore, it was of interest to determine whether any of those metabolites identified by PLS-DA with high VIP scores where statistically significant over time, as this may provide additional understanding and potential biomarker identification of this cohort of untargeted metabolites. When comparing to sedation-controls and correcting for Bonferonni adjustment for repeated measures (P-value <0.008), both hexose and p-hydroxybenzoate were significant at, at least one time point (Table 1).

**Table 1:** Repeated measures summary statistics for VIP ≥2.0 at baseline (A), 72 hours (B), and 8 days (C). Independent T-test performed (assuming equal variances); *F-test for equal variances was P = < 0.05 Welch-test (assuming unequal variances) was used; P-values less than 0.008 were deemed significant, after the Bonferonni adjustment; comparing MODS and ECMO samples to sedation-controls; TCDCA: tauro-chenodeoxycholic acid.

| Metabolites | Hydroxyphenylacetic acid |  |  |  | 2,3_dihydroxybenzoic acid |  |  |  | 2-keto-isovalerate |  |  |  | Deoxyribose phosphate |  |  |  | Hexose |  |  |  | p-hydroxybenzoate |  |  |  | TCDCA |  |  |  |
| --- | --- | --- | --- | --- | --- | --- | --- | --- | --- | --- | --- | --- | --- | --- | --- | --- | --- | --- | --- | --- | --- | --- | --- | --- | --- | --- | --- | --- |
|  | Mean | SD | Median | P-value | Mean | SD | Median | P-value | Mean | SD | Median | P-value | Mean | SD | Median | P-value | Mean | SD | Median | P-value | Mean | SD | Median | P-value | Mean | SD | Median | P-value |
| Sedation (n=4) | 47.40 | 53.93 | 44.87 |  | 16.00 | 10.56 | 13.58 |  | 6.52 | 0.97 | 6.63 |  | 6.79 | 4.60 | 6.71 |  | 15.80 | 3.26 | 14.37 |  | 53.12 | 7.66 | 53.00 |  | 0.22 | 0.323 | 0.08 |  |
| MODS BL (n=16) | 1.82 | 5.29 | 0.43 | P = 0.1900 | 1.81 | 2.63 | 0.74 | P = 0.0760 | 11.83 | 8.64 | 10.03 | P = 0.0291 | 1.09 | 1.12 | 0.73 | P = 0.0907 | 37.22 | 24.03 | 30.55 | P = 0.0031 | 65.01 | 36.71 | 56.51 | P = 0.2476 | 2.53 | 4.14 | 0.77 | P = 0.0422 |
| ECMO BL (n=8) | 4.72 | 10.12 | 0.81 | P = 0.2146 | 10.98 | 14.87 | 4.80 | P = 0.5631 | 9.33 | 3.91 | 9.41 | P = 0.0879 | 2.25 | 3.25 | 1.15 | P = 0.0733 | 21.03 | 19.43 | 23.88 | P = 0.4799 | 26.59 | 20.68 | 15.73 | P = 0.3170 | 14.30 | 25.76 | 2.53 | P = 0.1660 |
| MODS 72h (n=15) | 6.94 | 12.2 | 2.39 | P = 0.2331 | 2.10 | 3.15 | 1.23 | P = 0.0804 | 10.60 | 5.07 | 8.18 | P = 0.0127 | 0.79 | 0.65 | 0.62 | P = 0.0802 | 9.79 | 12.38 | 0.19 | P = 0.1228 | 19.16 | 15.29 | 13.74 | P = 0.0251 | 11.87 | 24.91 | 1.93 | P = 0.1037 |
| ECMO 72h (n=7) | 7.87 | 7.65 | 6.7 | P = 0.2410 | 5.12 | 4.39 | 4.32 | P = 0.0366 | 9.78 | 4.33 | 7.91 | P = 0.0981 | 1.40 | 0.77 | 1.31 | P = 0.1025 | 2.94 | 6.76 | 0.10 | P = 0.0065 | 41.73 | 20.51 | 39.74 | P = 0.0482 | 9.20 | 9.10 | 4.95 | P = 0.0402 |
| MODS 8d (n=8) | 24.04 | 19.50 | 20.97 | P = 0.4629 | 2.54 | 1.85 | 1.38 | P = 0.0855 | 14.04 | 6.96 | 11.71 | P = 0.0172 | 0.53 | 0.57 | 0.27 | P = 0.0729 | 2.77 | 7.46 | 0.14 | P = 0.0083 | 24.57 | 23.82 | 14.20 | P = 0.0021 | 792.38 | 2208 | 8.05 | P = 0.3441 |
| ECMO 8d (n=6) | 13.66 | 12.16 | 10.70 | P = 0.3062 | 4.68 | 7.96 | 1.45 | P = 0.0878 | 9.70 | 5.10 | 7.87 | P = 0.1874 | 0.77 | 0.34 | 0.68 | P = 0.0793 | 0.08 | 0.03 | 0.07 | P = 0.0024 | 9.05 | 3.047 | 9.20 | P < 0.0001 | 6.66 | 7.604 | 4.15 | P = 0.0929 |

Box and whisker-plots were generated to be able to visualize the distribution of samples over time (Figure 4). From this we can visualize a large spread of patient blood plasma metabolite values, and that the values change over the three time points. More frequent and intensive sampling would be necessary to determine exact distribution for the acute, stabilization and recovery phases of MODS and ECMO patients, using a metabolic platform. This illustrates the complexity and the dynamic nature of sampling for this patient population.

**Figure 4:**
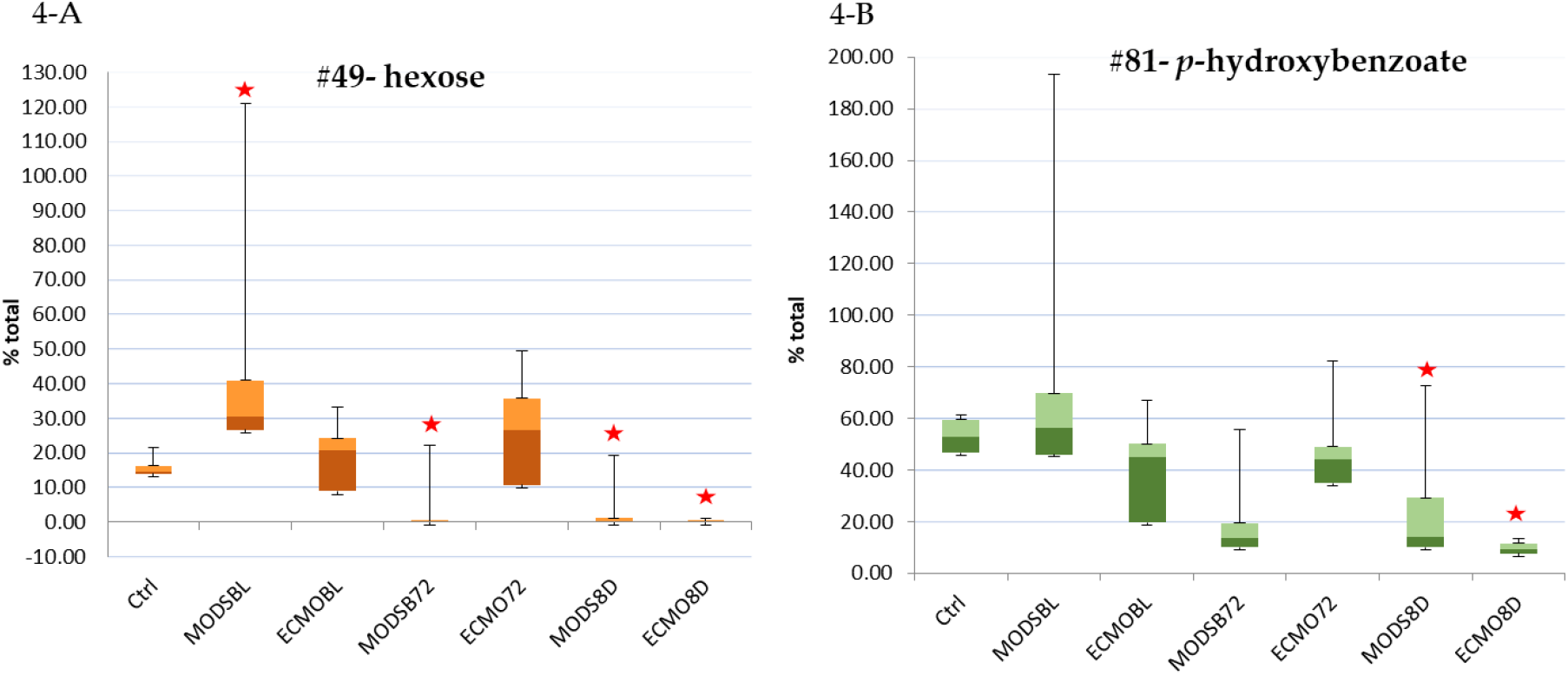
Top two metabolites with significant differences from sedation-controls over 8-days. Box and whisker plots, which include the median, the interquartile range (box), the outer range (whiskers) and pictorially summarize the central tendency, dispersion, skewness, and extremes of the dataset using a linear scale. MODS: multi-organ dysfunction syndrome; ECMO: extracorporeal membrane oxygenation; BL: baseline; 72: 72-hour time point; 8D: 8-day time point. Stars denote statistical significance from Independent T-test, or Welch-test as compared to sedation-controls.

### 3.4 Internal Validation

Kendall’s tau-b test was used for internal validation, and revealed a positive correlation between creatinine and the respective creatinine metabolite at all three time points (baseline: τb=0.708, p=0.000; 72 hours: τb=0.511, p=0.001; 8 days: 0.684, p=0.001) (Supplemental Figure 3). While the three coefficients yielded similar results, the skewness and heteroskedasticity of the data violate the assumptions for Pearson’s r. Spearman’s Rho and Kendall’s Tau are both non-parametric and acceptable to use in this case. Spearman’s Rho measures the rank correlation (how the ranks of the x and y values align), and Kendall’s Tau measures the percent of concordant pairs, which is also based on ranks but considered to be a more robust measure. Creatinine was found to be high at baseline, which correlated with clinical creatinine values.

## 4. Discussion

Untargeted analyses are intended to provide a means for finding differences in abundance across samples or groups of samples. Multivariate statistical techniques are therefore well suited to analysis of untargeted metabolomics data. MetaboAnalyst is a web-based platform created and developed by the Wishart group from the University of Alberta, offering R-based software for public use [20,26], and was the software of choice for this analysis.

From these preliminary results we may glean a few main findings: 1) metabolites of MODS and ECMO patients contain both Phase I and Phase II metabolites; 2) are dynamic in nature; 3) contain both potentially clinically relevant findings (such as TCDCA), as well as those of benign (±inert) function, as normally metabolized and excreted through downstream organ systems (renal, digestive); 4) metabolites can be measured and qualified in blood plasma of critically ill pediatric patients using an untargeted approach, which correlated to clinical values for routine care (e.g. creatinine).

Phase I and Phase II metabolites were detected amongst the 66 values analyzed. Recent evidence suggests that gut microbiome influences blood metabolites [27], which undergo Phase I metabolism through oxidation (R-OH), reduction (R-SH), or hydrolysis (R-NH_2_), and Phase II metabolism through sulfation (R-SO_3_H), glucuronidation (R-Gluc) and glutathione conjugation (R- Gl). This may suggest that there are greater metabolic influences beyond the scope this this study that need to be controlled for in future work, such as including gut microbiome profiles.

*p*-hydroxybenzoate, is thought to be produced via two major pathways: 1) microbial oxidation of petroleum derivative toluene into *p*-hydroxybenzoate, as described for *Pseudomonas* species [28]; and 2) the *de novo* bioproduction of *p*-hydroxybenzoate from amino acids (tyrosine, tryptophan, phenylalanine) through and intermediate chorismite, via the enzyme chorismite lyase (UbiC) [29]. *p*-hydroxybenzoate (paraben and alkyl ester derivative) is commercially used as a preservative and antimicrobial agent pharmaceutical and cosmetic industry [30], and therefore may be from an exogenous source. The second possibility is an endogenous source, which has been described in *Escherichia coli* [31] and *Mycobacterium tuberculosis* [32], as produced from glucose, and can be toxic at higher-concentrations. Lower levels of *p*-hydroxybenzoate as observed in our MODS and ECMO patients at day 8 is a Phase I metabolite, and may be an indication of a gut bacteria dysbiosis (impaired microbiota), and to date has not been described in this patient population.

The most dynamic metabolite described herein is hexose which reflects the fluctuating energetic state of the patients. Low hexose at the third time point (8 days-post MODS diagnosis and into their PICU admission) could be a sign of energy deficiency, however we know that the patients were under close monitoring and by 72 hours were all receiving some nutritional intervention [12], after largely being *nil per os* at baseline. Blood glucose control has been further evaluated in Covid-19 patients, and is of ongoing concern given reports of higher mortality and multi-organ injury [33]. HPLA, a phenylcarboxylic acid, has been speculated to be a marker of sepsis in adult cardiac surgery, however requires further validation [34].

Cholesterol breaks down in the liver to produce primary bile acids, one being chenodeoxycholic acid, which together with taurocholic acid produces a conjugated bile acid TCDCA and excreted in the intestine, constituting our enterohepatic circulation. In spite of normalizing liver enzymes (alanine transaminase (ALT) Aspartate transaminase (AST)) in this patient population as previously reported [11], TCDCA remained elevated as compared to sedation-controls at the 8-day time point. This indicates that the metabolic profile may illustrate a different landscape on patient recovery, and metabolites may be organ specific.

Limitations of the work include a low sample volume, capturing high-abundance metabolites, and sample integrity may have been compromised by a previous freeze-thaw cycle. A second cohort study would be necessary for further metabolite identification and validation to explore their clinical utility.

## 5. Conclusions

It is feasible to measure blood plasma metabolites in pediatric patients with MODS and undergoing ECMO treatment. Metabolites may be benign or significant in describing a patient’s pathophysiology, fluctuate over time and require operator interpretation.

## Data Availability

The data is available upon request.

## 6. Abbreviations

HPLA: hyroxyphenylacetic acid
LC-MS: Liquid chromatography-mass spectrometry
MODS: multi-organ dysfunction syndrome
PICU: pediatric intensive care unit
PLS-DA: partial least squares-discriminant analysis
VA-ECMO: veno-atrial extracorporeal membrane oxygenation
VIP: variance of importance
TCDCA: tauro-chenodeoxycholic acid.

## Supplementary Materials

Table S1: Metabolites identified n=115, metMz-*mass over charge ratio*, metRt- *retention time*, compound name, and compound ID listed; Figure S1: a) Metabolic actions of 115 metabolites identified; b) Ontological sources of 115 metabolites identified, taken from Human Metabolite Data Base (HMDB) (https://hmdb.ca/metabolites/); Figure S2: Clustering result shown as heatmaps at baseline (A), 72 hours (B), and 8 days (C); Figure S3: Correlation of clinical creatinine to untargeted metabolite value(s).

## Author Contributions

SR, DG and ML conceived of the original study design, screened, recruited, and collected all samples. Sample processing, peak findings, annotation, and quantification for metabolites was performed by TL. ML and AK performed all statistical and bioinformatic analysis. CK contributed to the annotation of individual metabolites. SR, DG, ML, AK, CK, BP, DS, JP and TL all contributed to the writing and editing of the manuscript.

## Acknowledgments

The authors would like to thank the PICU staff at HDVCH for their support in the completion of this study and various contributions. Dr. David Tack and Dr. Alan Davis, for their additional statistical support.

