## Supplemental for "The Feasibility of Studying Metabolites in PICU Multi-Organ Dysfunction Syndrome Patients Over an 8-day Course Using An Untargeted Approach"

Article

Received: date; Accepted: date; Published: date

**Abstract:** Metabolites are generated from critical biological functions and metabolism. This pediatric study reviewed plasma metabolites in patients suffering from multi-organ dysfunction syndrome (MODS) in the pediatric intensive care unit (PICU) using an untargeted metabolomics approach. Patients meeting criteria for MODS were screened for eligibility and consented (n=24), and blood samples were collected at baseline, 72 hours, and 8 days; control patients (n=4), were presenting for routine sedation in an outpatient setting. A sub-set of MODS patients (n=8) required additional support with veno-atrial extracorporeal membrane oxygenation (VA-ECMO) therapy. Metabolites from thawed blood plasma were determined from ion pairing reversed-phase LC-MS analysis. Chromatographic peak alignment, identification, relative quantitation, statistical and bioinformatics evaluation were performed using MAVEN and MetaboAnalyst 4.0. Metabolite analysis revealed 115 peaks per sample. From the PLS-DA with VIP scores above ≥2.0, 7 dynamic metabolites emerged over the 3 time points: tauro-chenodeoxycholic acid (TCDCA), hexose, *p*-hydroxybenzoate, hydroxyphenylacetic acid (HPLA), 2_3-dihydroxybenzoic acid, 2-keto-isovalerate, and deoxyribose phosphate. After Bonferonni adjustment for repeated measures hexose and *p*-hydroxybenzoate were significant at one time point, or more. Kendall's tau-b test was used for internal validation of creatinine. Metabolites may be benign or significant in describing a patient’s pathophysiology and require operator interpretation.

**Supplementary Materials:**

**Table S1:** Metabolites identified n=115, metMz-*mass over charge*, metRt-*retention time*, compound name, and compound ID listed.

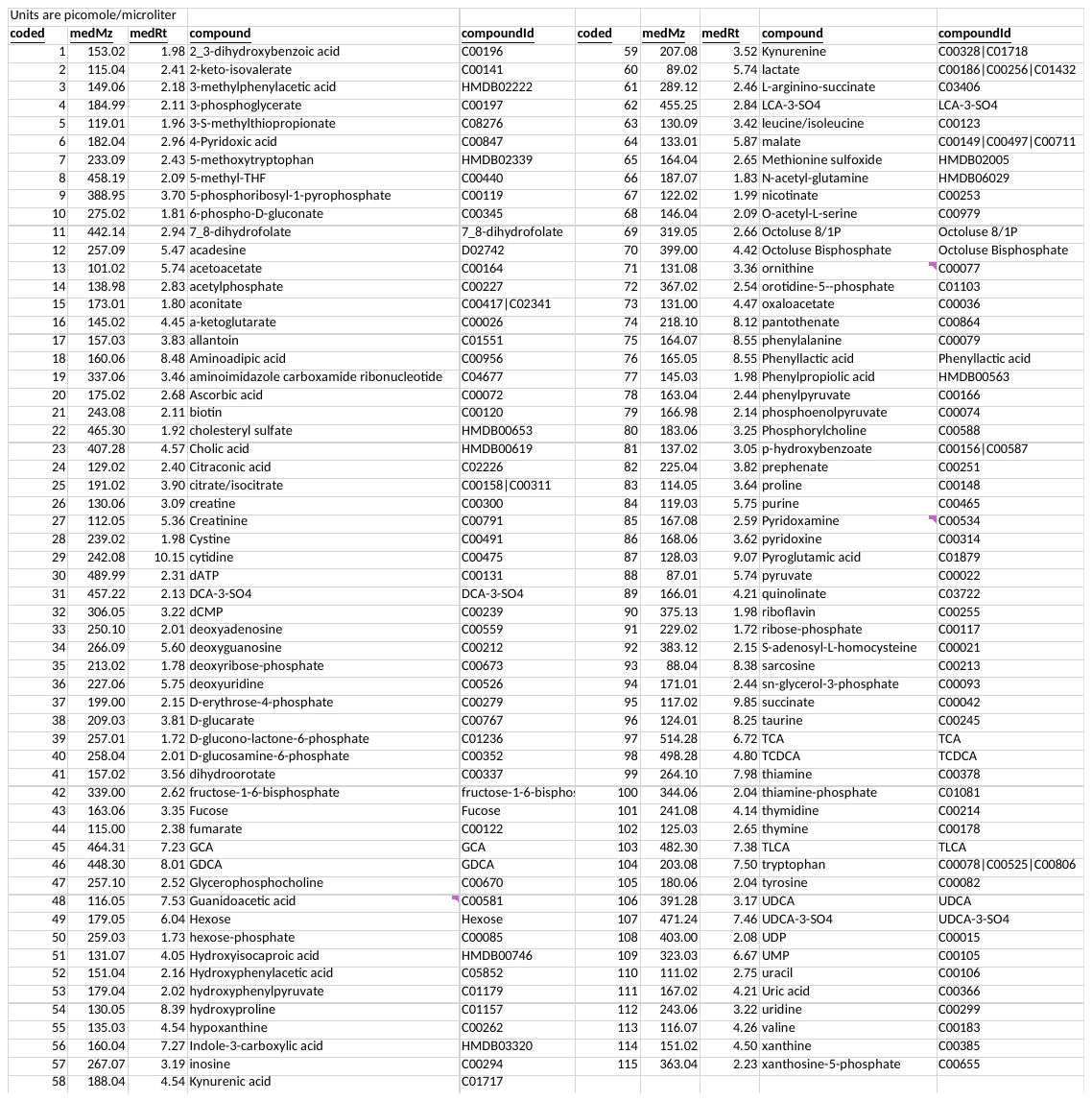

**Figure S1:** a) Metabolic actions of 115 metabolites identified; b) Ontological sources of 115 metabolites identified, taken from Human Metabolite Data Base (HMDB) (https://hmdb.ca/metabolites/).

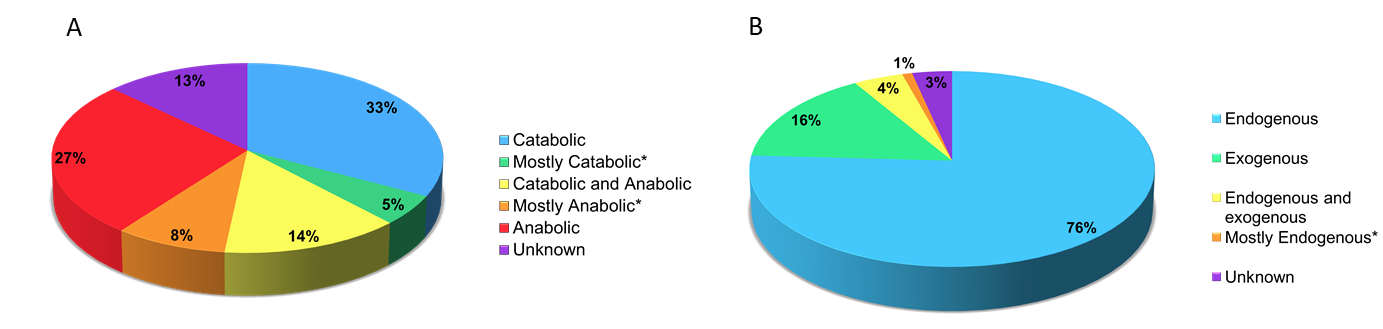

**Figure S2:** Clustering result shown as heatmaps at baseline (A), 72 hours (B), and 8 days (C).

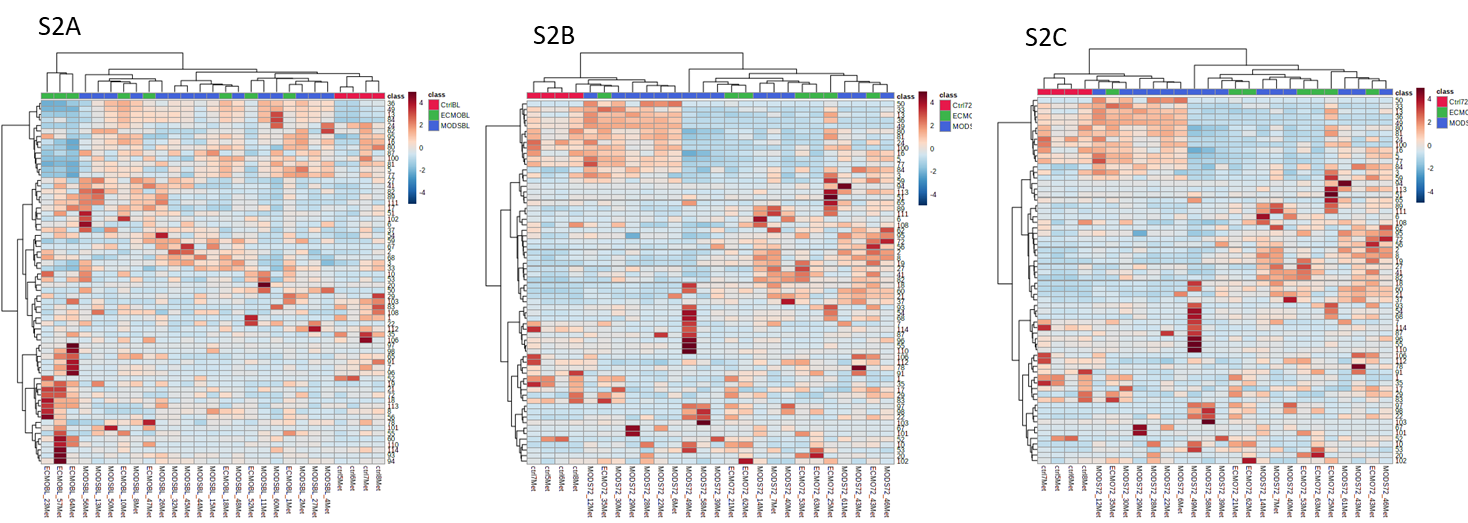

**Figure S3:** Correlation of clinical creatinine to untargeted metabolite value(s).

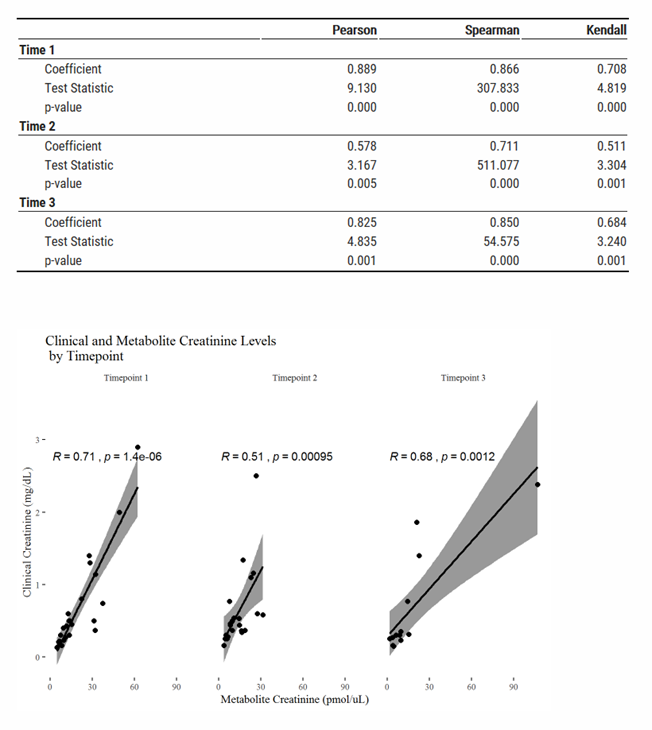
